# A comprehensive survey on the beliefs, perceptions, and clinical manifestations of pre and post Covid-19 vaccinations among physiotherapy students in the United Arab Emirates

**DOI:** 10.1101/2023.02.17.23285959

**Authors:** Shaikha Almheiri, Animesh Hazari, Praveen Kumar, Sampath Kumar, Srilatha Girish

**Author notes:** Corresponding Author: **Dr. Animesh Hazari (MS, Ph.D.), Assistant Professor, College of Health Sciences, Gulf Medical University, Email**-, **Ph- **. All authors have contributed equally to this work.

## Abstract

**Background:** The World Health Organization has defined Coronavirus Disease (COVID-19) as an infectious pandemic, caused by a newly discovered SARS-CoV-2 virus. Students relied heavily on the internet, social media, parents, and friends, in addition to medical advice for information on its presentation, complications, prevention, and management. It is evident from the literature that healthcare professionals including students who play an important role in the healthcare system may be lacking important information on COVID-19 vaccinations. Thus, the study aims to identify and compare the pre-post covid-19 vaccination-related essential information among Physical Therapy university students.

**Methods:** A cross-sectional survey was conducted among physiotherapy university students in the United Arab Emirates using a self-administrated structured questionnaire. The questionnaire comprised three sections covering beliefs, perceptions, and clinical manifestations of the pre and post-COVID-19 vaccination. The survey was shared with more than 300 students through email and social platforms during the time between January 2022 till December 2022.

**Results:** The majority of the physiotherapy students believed the vaccines to be safe (71.3%) due to multiple reasons while others did not believe in the effectiveness of the vaccine (28.7%). Some students did report unusual symptoms (painful periods, hair loss, forgetfulness) after the vaccine (10%). Similar findings were reported for family members as well (14%). Students had a positive perception of the vaccine and reported willingness to take it even if not mandatory (68%).

**Conclusion:** Some students did believe in the safety of the vaccine due to multiple reasons while some did not due to fear of long-term side effects and personal choices. The finding of the study could be useful to create confidence as well as awareness among physiotherapy students as they are often invited to aid during medical pandemics such as Covid 19. Also, higher rates of vaccination among healthcare professionals will impart higher acceptance in the medical organization due to safety guidelines. This could also help to counsel other students against fear and apprehension towards the vaccination of such kind in the future.

## Introduction

At the end of 2019, a new virus was discovered causing severe respiratory symptoms in Wuhan, China. The World health organization has defined it as coronavirus disease (Covid-19). The main method of transmission is through droplets of the infected person (through coughing, or sneezing) [1]. Covid-19 affects individuals in different ways, the majority of the reported cases did not require hospitalization [2]. The elderly and people with existing co-morbidities such as diabetes and cardiovascular disease are prone to develop serious illnesses [3]. First six months of the pandemic, people faced a lot of fear and hesitation to carry on with their daily life routines [4], where a lot of precautionary measures were implemented. Many vaccine trials were put into research and development to combat the situation but the hesitancy and anxiety towards vaccination continued due to various reasons.

By July 2020, the clinical trials of the Sinopharm vaccine were launched in the United Arab Emirates and efforts were made to vaccinate the mass for protection against the infectious disorder [5]. Later Pfizer and Sputnik V were introduced, and the vaccines were approved by the food and drug administration (FDA) in mid-2021 for emergency use since the expected benefits outweigh the risks and side effects[6]. In UAE Sinopharm, Pfizer, and Sputnik V were available to the public, and priority was given to the front-line workers and elderly. Slowly it was made available to all ages starting from 16 years. Once it was released, the vaccines faced a lot of refusals mainly due to misinformation spreading through social media leading to vaccine hesitations [7]. National campaigns were started to encourage people to take the vaccine which was successfully leading to 97% of the population being fully vaccinated as per the statistics from the UAE national emergency crisis and disasters management authority [8]. The willingness to get vaccinated and the factors associated with was a major area of research globally and the reports varied across the globe. A study conducted in Kuwait reported that 53.1 percent of the population was willing to get full vaccination once the vaccines were available [9]. A similar study conducted in China reported a willingness rate of 79.08% among the elderly compared to 85.75 in adults [10]. In the middle east, a study conducted in Jordon reported a willingness rate of 36.8% and 26.4% rejected it suggesting a significantly higher rate of hesitancy [11]. However, no data was available on the willingness and acceptance of vaccinations among the different populations as well as age groups in the UAE. It was hypothesized that a higher vaccination rate as reported by the local government portal could be adopted as per strict government norms and not reflect the actual willingness towards vaccination. Understanding the willingness or hesitancy could generate higher acceptance as well as help medical professionals to perform their duty efficiently. Medical students are among the group of frontline healthcare providers likely to be exposed to Covid-19 patients. Among various medical professionals, physical therapy has an important contribution as the front-line healthcare workers, and their beliefs and perception would have a significant role in driving the outcome of Covid-19. University students with a practicing physical therapy license were invited as an additional task force to handle the pandemic across the world. In UAE as well when the necessity arose, university medical students including physiotherapy students were called for assistance in the management of Covid-19. However, there was a significant disparity in their beliefs, perception, and knowledge of clinical manifestation towards Covid 19 vaccination which hindered their understanding of the disease and ultimately affected their management skills. It was important to achieve high Covid-19 vaccination coverage rates in this group as soon as a vaccine was available.

With the review of the literature, we found that there was a major dearth of data for belief, perception, and knowledge among university medical students. In UAE, a study published in January 2022 targeting university students, in general, reported a vaccination rate of 38.8% with a sample size of 467 students [12]. However professional stratification has not been reported which could vary due to differences in exposure, knowledge, and prior experience.

Thus, the study aimed to identify and compare the pre-post Covid-19 vaccination-related essential information among physical therapy university students. The information was required to fill the gaps in understanding Covid-19 and help physical therapists efficiently manage the condition with scientific rigor. Moreover, being a medical professional, a physical therapist may have a different understanding of the disorder which might be reflected through their belief and perception and thus should be explored [13]. The objectives of the study were as follows:

1. To identify the post-Covid-19 vaccination-related clinical manifestation in physiotherapy students.
2. To assess the perception and beliefs about the Covid-19 vaccine in physiotherapy students pre and post-vaccination.

## Methodology

### Research design and setting

A cross-sectional, web-based survey study was conducted at the Department of Physical Therapy, College of Health Sciences, Gulf Medical University, United Arab Emirates.

### Study population

All students enrolled in bachelor and master programs for Physical Therapy were invited through google forms to participating in the study. There are three universities currently offering the Physical Therapy program in UAE (Gulf Medical University Ajman, University of Sharjah, Sharjah, and Fatima College of Health Sciences, Al Ain). A total of approximately 500 students are currently enrolled in the bachelor and master programs across three institutions. The calculated sample size was 218 (95% confidence interval, a margin of error= 0.05) [14]. The survey was sent to all students. The response rate was initially low and multiple reminders and requests were sent through all possible communication. The participants were recruited as per the given inclusion and exclusion criteria given below.

### Inclusion criteria

- all physiotherapy students actively enrolled in bachelor and master program
- students who completed two doses of Vaccines recognized by the UAE govt.
- the second dose should have been completed within the past six months
- students of any nationality studying at selected universities in UAE.

### Exclusion criteria

- students of any other health profession
- who didn’t give consent
- active Covid-19 infection

### Study procedure

The study was approved by the Institutional Review Board, College of Health Sciences, Gulf Medical University (Ref no-IRB/COHS/STD/07/Jan-2022). A structured questionnaire was prepared which comprised three sections covering beliefs, perceptions, and clinical manifestations of the pre-post Covid-19 vaccine. A set of 30 questions was initially prepared and presented to the experts (medical doctor, physiotherapist, psychologist) to validate, after the feedback from the experts, 8 questions were excluded upon disagreement and 22 questions were taken into the next phase. In the 2^nd^ phase, the questionnaire was reshared with the experts, and the validity of the questions was tested over a descriptive scale as given below:

**4:** highly significant (to be included)

**3:** significant (to be included)

**2:** less significant (to be excluded)

**1:** not significant (to be excluded).

All reviewers scored the individual questions and the questions with the highest scores in order were included in the final questionnaire (15 out of 22). The survey was created through google forms for each section (belief, clinical manifestation, and perception). Answers options included a “Yes or No” rating on a scale (high to low, increased to decreased), and a list of multiple options that could be selected with the option of “other”. Informed consent was taken from all participants before filling in the responses to the questionnaire. If the participant declined to participate, exclusion was made, and the record was removed from the data analysis procedure. The questionnaire was shared through email and randomly distributed among physiotherapy students in UAE. The study was conducted from January 2022-December 2022.

### Statistical analysis and data acquisition

SPSS V21 was used for data analysis. The questionnaire responses were coded for descriptive analysis and the results have been presented in the tables below.

## Results

The important findings of the study have been presented in Table 1-Table 5.

**Table 1.** Demographic data of all participants

| Demographic data (n:150) |  |  |
| --- | --- | --- |
| Gender |  |  |
|  | Frequency | Percent |
| Male | 40 | 26.6 |
| Female | 110 | 73.3 |
| Age |  |  |
| 18-20 | 67 | 44.6 |
| 20-25 | 50 | 33.3 |
| 25-30 | 27 | 18 |
| >30 | 6 | 4 |
| College Program |  |  |
| Bachelor of physical therapy | 117 | 78 |
| Master of physical therapy | 33 | 22 |

## Discussion

In the given study. a total of 174 participants responded to the survey. However, 24 responses were incomplete, therefore we took 150 completed responses for data analysis as per the study timeline making a responder rate of 70%. Table 1 represents the demographic characteristics of the participants where the participation was dominated by females comprising 110 of 150 samples. The majority of the participants were within the age group of 18-25 years and six participants were above 30. These findings are suggestive of a homogenous population and the responses could be considered a direct representation of the sample population. The higher number of female participants could be attributed to the professional academic choice towards the medical stream. Our study findings suggested that there was uniformity among the sample population towards the selection of different types of vaccines. Fifty-three (35.3%) students had taken Sinopharm, while forty-five (30%) took Pfizer and fifty-two (34.7%) have taken both Sinopharm and Pfizer (Table 2). However, it should be noted that in UAE, Sinopharm was first available and later Pfizer was administered as a single and booster dose. It was also observed that people who took Sinopharm also took Pfizer as a booster dose which may reflect their confidence in Pfizer more compared to Sinopharm.

**Table 2:**
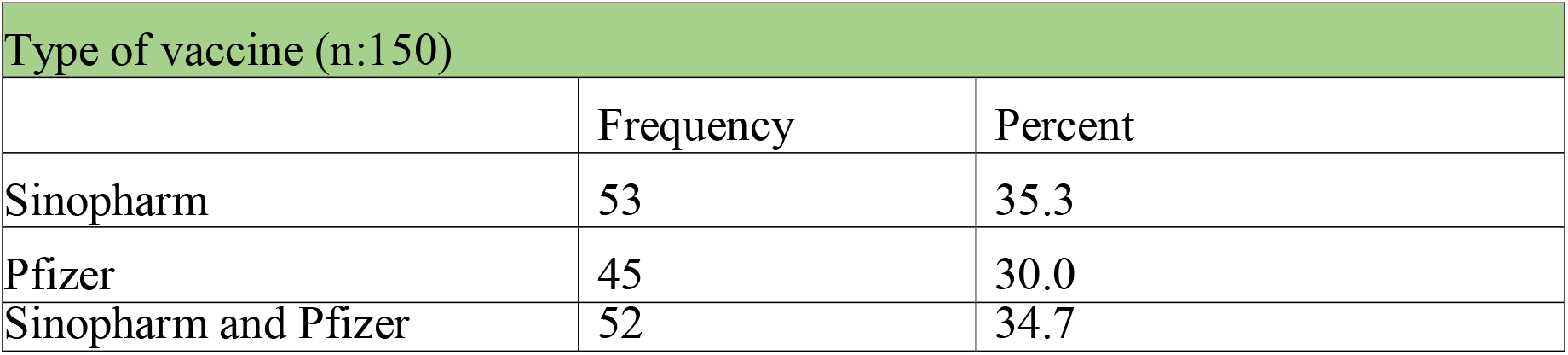
Type of vaccines administered

In context to the responses towards “beliefs” (Table 3), 107 (71.3%) students believed that the vaccines were safe while 43 (28.7%) believed otherwise. One of the most common reasons for the students to believe that they are safe was associated with the safety ensured by the World Health Organization (WHO) with a frequency of 27.3% (n=41). It indicates the importance and influence the WHO has on the community and the younger age group. The second most common finding was attributed to scientific knowledge where the students trusted the effectiveness of the vaccine due to the research papers being published and supporting the vaccine (39 comprising 26% of the population). Hence, it reveals that the current generation of students does follow up with recent publications while making decisions on a scientific basis. Lastly, 28 (18.7%) trusted the vaccine safety because several people whom they know have taken the vaccine and they were safe. Therefore, peer reviews also play an important role. Participants who responded to the unsafety of the vaccines believed that it could cause a variety of concerns. In this context, the students were asked to choose multiple options of what they thought the vaccine can cause. Forty-five (30.0%) believed that the vaccine could cause headaches, fever, and body aches while 24 (16.0%) believed it can cause headaches and fever only. Interestingly, on the other hand, a small number of students did believe that it can cause infertility 2 (1.3%) and paralysis and cancer 3(2.0%). The reasons for such a belief and fear could not be assessed due to personal reasons, but it was clear that they failed to justify it with a scientific reason and there could be a stigma associated with its acceptance even in professionals from a healthcare background which could be alarming and requires further investigation. Rating the fear of getting the Covid-19 infection before the vaccine, 46 (30.7%) reported low levels of fear, 68 (45.3%) had moderate fear and 36 (24%) confronted high fear. While after receiving the vaccine, 79(52.7%) reported reduced levels of fear, 5 (3.3%) had increased levels and 66 (44.0%) reported their fear is still the same. The number for low fear increased post-vaccination which was a positive sign. However, there was a greater cohort of students who believed their fear levels were the same. The reasons for such findings were reported as not believing in the effectiveness of the vaccine in addition to believing that there their immunity was low.

**Table 3:** Responses related to “Belief”

| Belief |  |  |
| --- | --- | --- |
| Do you believe that covid 19 vaccines are safe |  |  |
|  | Frequency | Percent |
| Safe | 107 | 71.3 |
| Not safe | 43 | 28.7 |
| What makes you believe that they are safe |  |  |
| Safety ensured by WHO | 41 | 27.3 |
| Publications supporting the effectiveness | 39 | 26.0 |
| Several people have taken it and are safe | 28 | 18.7 |
| I believe Covid 19 vaccine can cause |  |  |
| Infertility | 2 | 1.3 |
| headaches | 5 | 3.3 |
| Fever | 12 | 8.0 |
| body aches | 3 | 2.0 |
| Cancer | 2 | 1.3 |
| Infertility, paralysis | 3 | 2.0 |
| Infertility, body aches | 1 | 0.7 |
| Infertility, cancer | 1 | 0.7 |
| Paralysis, headaches | 1 | 0.7 |
| Paralysis, cancer | 3 | 2.0 |
| Headaches, fever | 24 | 16.0 |
| Headaches, body aches | 2 | 1.3 |
| Headaches, cancer | 1 | 0.7 |
| Fever, body aches | 13 | 8.7 |
| Cancer and forgetfulness | 2 | 1.3 |
| Paralysis, headaches, fever, body aches | 3 | 2.0 |
| Headaches, fever, body aches | 45 | 30.0 |
| Infertility, headaches, fever, body aches | 1 | 0.7 |
| Infertility, paralysis, fever, body aches, forgetfulness | 1 | 0.7 |
| Infertility, fever, body aches | 1 | 0.7 |
| Infertility, headaches, fever | 1 | 0.7 |
| Paralysis,headaches,<br>bodyaches, forgetfulness | 1 | 0.7 |
| Headaches, fever, bodyaches,<br>forgetfulness | 2 | 1.3 |
| Paralysis ,headaches,fever | 1 | 0.7 |
| Headaches, fever,bodyaches ,<br>cancer, forgetfulness. | 1 | 0.7 |
| Infertility, cancer,<br>forgetfulness | 1 | 0.7 |
| Infertility, paralysis,<br>headaches, fever, body<br>aches, cancer,<br>forgetfulness | 2 | 1.3 |
| Infertility, paralysisheadaches | 1 | 0.7 |
| Infertility,fever,bodyaches,<br>cancer | 1 | 0.7 |
| Bodyaches,cancer,<br>forgetfulness | 1 | 0.7 |
| Paralysis, bodyaches,cancer,<br>forgetfulness | 1 | 0.7 |
| Infertility, paralysis ,<br>cancer | 2 | 1.3 |
| Infertility, paralysis,<br>headaches, fever | 1 | 0.7 |
| Infertility, headaches,fever,<br>body aches | 1 | 0.7 |
| Infertility, headaches,fever,<br>forgetfulness | 1 | 0.7 |
| Infertility, headaches, fever,<br>body aches, forgetfulness ,<br>death, septic shock , organ<br>failure, respiratory<br>defects | 1 | 0.7 |
| Infertility, paralysis,headaches,<br>fever, bodyaches | 1 | 0.7 |
| Headaches, fever, bodyaches,<br>cancer | 1 | 0.7 |
| No one knows | 1 | 0.7 |
| Infertility,<br>paralysis, cancer,<br>forgetfulness | 1 | 0.7 |
| Fever, bodyaches,<br>cancer, forgetfulness | 1 | 0.7 |
| Rate your fear of getting covid 19 infection |  |  |
| Pre vaccine |  |  |
| Low | 46 | 30.7 |
| Moderate | 68 | 45.3 |
| high | 36 | 24.0 |
| Post vaccine |  |  |
| Reduced | 79 | 52.7 |
| Increased | 5 | 3.3 |
| Same | 66 | 44.0 |
| If same, why |  |  |
| I don't belief in<br>effectiveness of vaccine | 53 | 35.3 |
| I beliefs my immunity is<br>low | 25 | 16.7 |
| Reducing preventive measures with increasing rate of vaccinated people |  |  |
| Yes | 84 | 56.0 |
| No | 66 | 44.0 |

Regarding reducing the preventive measures with increasing rates of vaccinated people, such as wearing masks and maintaining social distancing, 84 participants (56%) chose “Yes” and 66 (44%) were against it, thus making a selection for “No”.

A higher number of responders with “ No” suggested that people could still believe that the vaccine alone may not be secure to prevent infection.

Considering the responses on “Perception”, Table 4 represents the important findings from the study. Responses were obtained for rating physical activity levels pre and post covid 19 vaccines. Findings suggested that pre-vaccine 31 students (20.7%) had low levels of physical activity, 106 (70.7%) moderate, and 13 (8.7) high. Post-vaccine 101(67.3%) had the same level as pre, whereas 40 (26.7%) responded to decreased levels and 9 (6%) to increased levels of physical activity. It was surprising to see that physical activity dropped post-vaccination significantly suggesting that physiotherapy students perceived exercise to affect their regular daily routine negatively. In an ideal situation, physical activity should have increased among healthcare professionals such as physiotherapists since they understand the importance of active life much better but the reduction in prior levels of physical activity post-vaccination was a clear sign of poor physical and mental health as a consequence of vaccination. We tried to figure out the possible reasons for the same. A list of reasons why they experienced the decreased levels was: 11 students (7.3%) experienced increased fatiguability, 6(4%) increased breathless and reduced aerobic capacity, and 6 (4%) reported increased fatiguability with joint and muscle. Other reasons such as general pain and less common reasons are mentioned in Table 4. In addition, in other domains of perception such as if the vaccines could trigger autoimmune disorders in individuals with a positive family history, a higher percentage answered “Yes” (86 (57.3%)), and 64 (42.7%) responded “No”. In January 2022, a study was published with results indicating the new onset of autoimmune manifestations such as autoimmune liver diseases, Guillen barre syndrome, and IgA nephropathy associated with covid 19 vaccine [15]. The findings of our study agreed with previous findings regarding the perception of autoimmune disorders related to Covid 19 vaccines. In addition, our study reported a significant number of participants who perceived forgetfulness and confusion post-vaccination (48,32%). A case study reported vaccine-induced immune thrombotic thrombocytopenia which led to confusion in a 63-year-old female patient [16]. A similar case study published in 2022 explored the possibility of cognitive deficits post covid 19 vaccines, and it concluded that the onset of acute cognitive deficits and memory impairments (psychosis) can be a complication after the vaccine and it requires further study by neurologists and physicians. [17]. A study has reported that the most common neurological symptoms post covid vaccines include dizziness, headache, pain, muscle spasms, myalgia, and paresthesia [18]. On the contrary, a systematic review reported evidence-based minor risk for acute neurological disorders post covid vaccination [19]. It should be noted that are several studies that report cognitive impairments after Covid-19 infection. A study has reported cognitive deficits such as Long Covid-19 complications [20]. Therefore, it is possible that the cognitive symptoms as reported in our study could overlap and partially be seen as a consequence of covid 19 infection and not directly as consequences of Covid-19 vaccination. The perception that Covid-19 vaccines could lead to forgetfulness and confusion could be a masked area of further research as there is a dearth of literature. Findings based on case reports cannot generate strong evidence and are incomparable to our study population.

**Table 4:** Responses related to “Perception”

| Perception |  |  |
| --- | --- | --- |
| Rate your physical activity levels pre and post Covid 19 vaccines |  |  |
| Pre vaccine |  |  |
|  | Frequency | Percent |
| Low | 31 | 20.7 |
| Moderate | 106 | 70.7 |
| High | 13 | 8.7 |
| Post vaccine |  |  |
| Same | 101 | 67.3 |
| Decreased | 40 | 26.7 |
| Increased | 9 | 6.0 |
| If decreased, what are the reasons |  |  |
| Increased breathlessness | 5 | 3.3 |
| Reduced aerobic capacity | 3 | 2.0 |
| Increased fatiguability | 11 | 7.3 |
| Joint and muscle pain | 5 | 3.3 |
| Increased breathlessness, increased fatiguability, reduced aerobic capacity, joint and muscle pain | 2 | 1.3 |
| Increased breathlessness, increased fatiguability, reduced aerobic capacity | 2 | 1.3 |
| Increased breathlessness, increased fatiguability | 4 | 2.7 |
| Increased breathlessness, reduced aerobic capacity | 6 | 4.0 |
| Increased fatiguability, joint and muscle pain | 6 | 4.0 |
| Increased breathlessness, increased fatiguability, joint and muscle pain | 1 | 0.7 |
| Reduced aerobic capacity , increased fatiguability | 1 | 0.7 |
| Increased breathlessness, joint and muscle pain | 1 | 0.7 |
| Reduced aerobic capacity, joint and muscle pain | 1 | 0.7 |
| I perceive that covid 19 vaccine can trigger autoimmune disorders in individuals with a history |  |  |
| Yes | 86 | 57.3 |
| No | 64 | 42.7 |
| Do you perceive forgetfulness and confusions post covid 19 vaccine |  |  |
| Yes | 48 | 32.0 |
| No | 102 | 68.0 |
| If vaccine was not a mandatory requirement to attend class, would you take it |  |  |
| Yes | 102 | 68.0 |
| No | 48 | 32.0 |
| If no, please mention your reasons |  |  |
| Need more time to see effectiveness | 6 | 4.0 |
| Not enough research | 3 | 2.0 |
| Unnecessary | 5 | 3.3 |
| Concerns about effectiveness | 5 | 3.3 |
| Fear of needles | 1 | 0.7 |
| Waste of time | 1 | 0.7 |
| It is new | 4 | 2.7 |
| We don't know what it is inside | 3 | 2.0 |
| Dangerous | 2 | 1.3 |
| Long term effects | 3 | 2.0 |
| Personal choice | 5 | 3.3 |
| Does not make a difference | 1 | 0.7 |
| I prefer not to take it | 1 | 0.7 |
| I don't believe in the effectiveness | 2 | 1.3 |
| I know people who died from vaccine | 1 | 0.7 |
| I was okay with weekly PCR; I did not want the vaccine | 1 | 0.7 |
| Covid 19 is based on immunity | 1 | 0.7 |
| I developed anxiety | 1 | 0.7 |
| Would you encourage your friends and family members to take the vaccine |  |  |
| Yes | 110 | 73.3 |
| No | 40 | 26.7 |

**Table 5:**
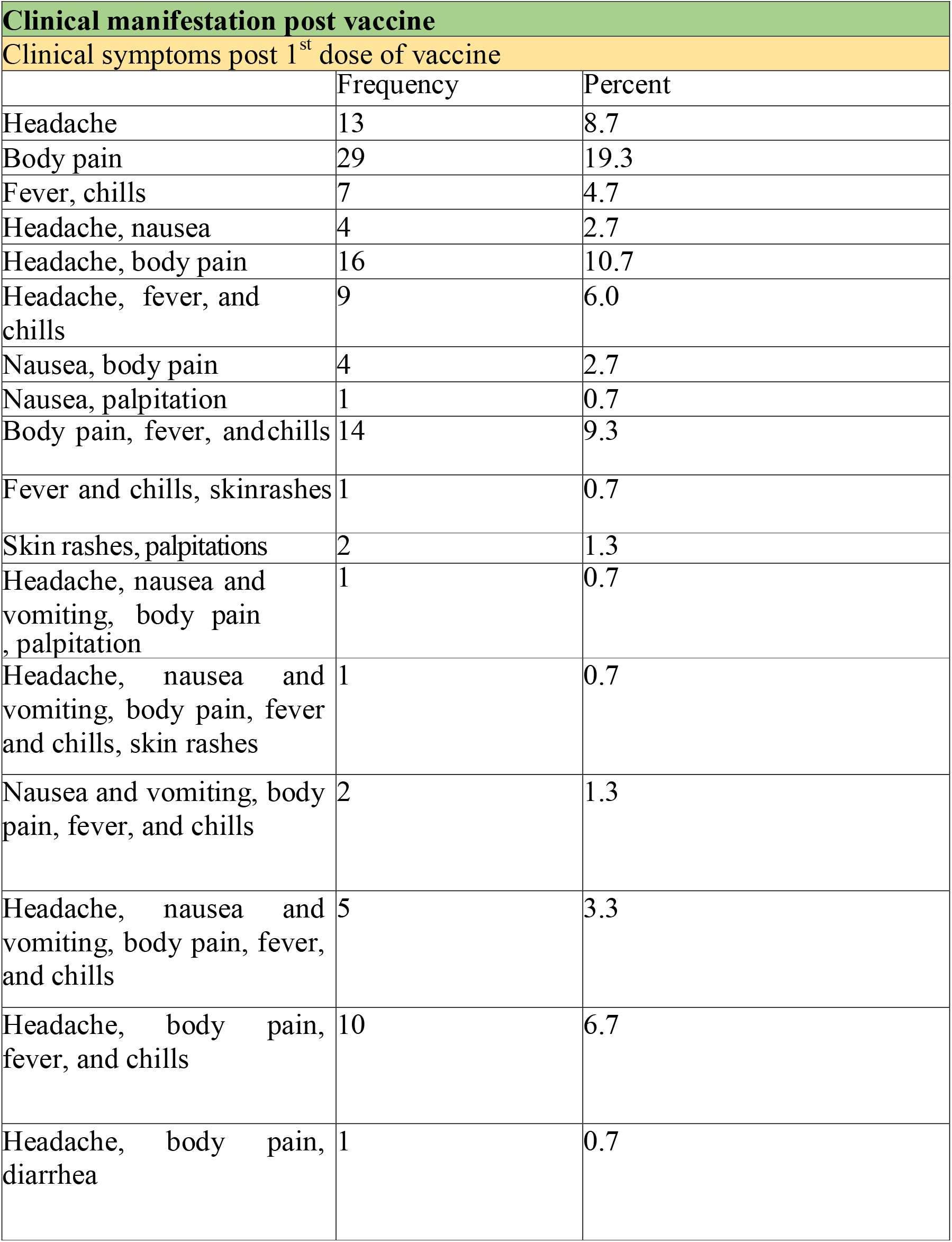
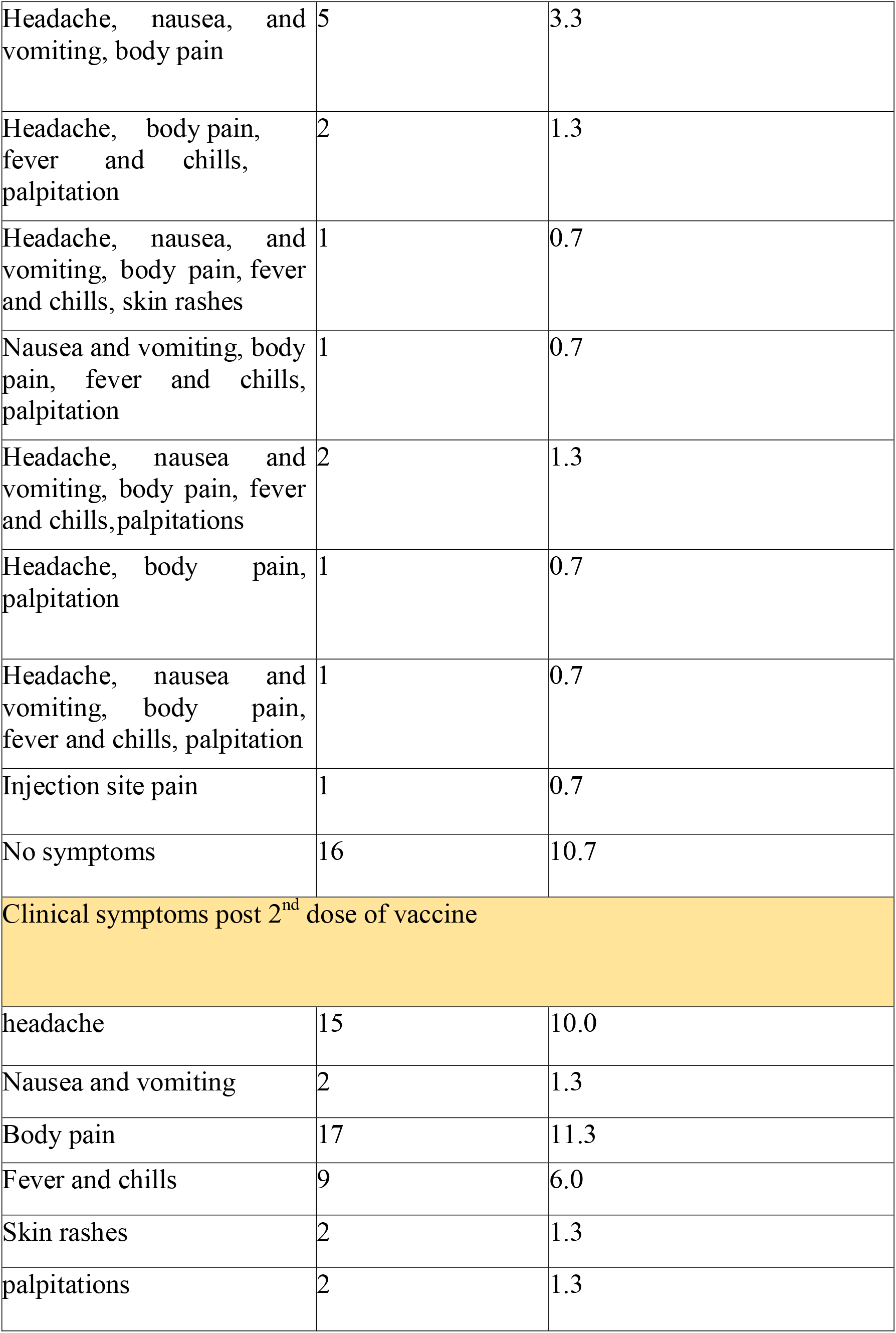
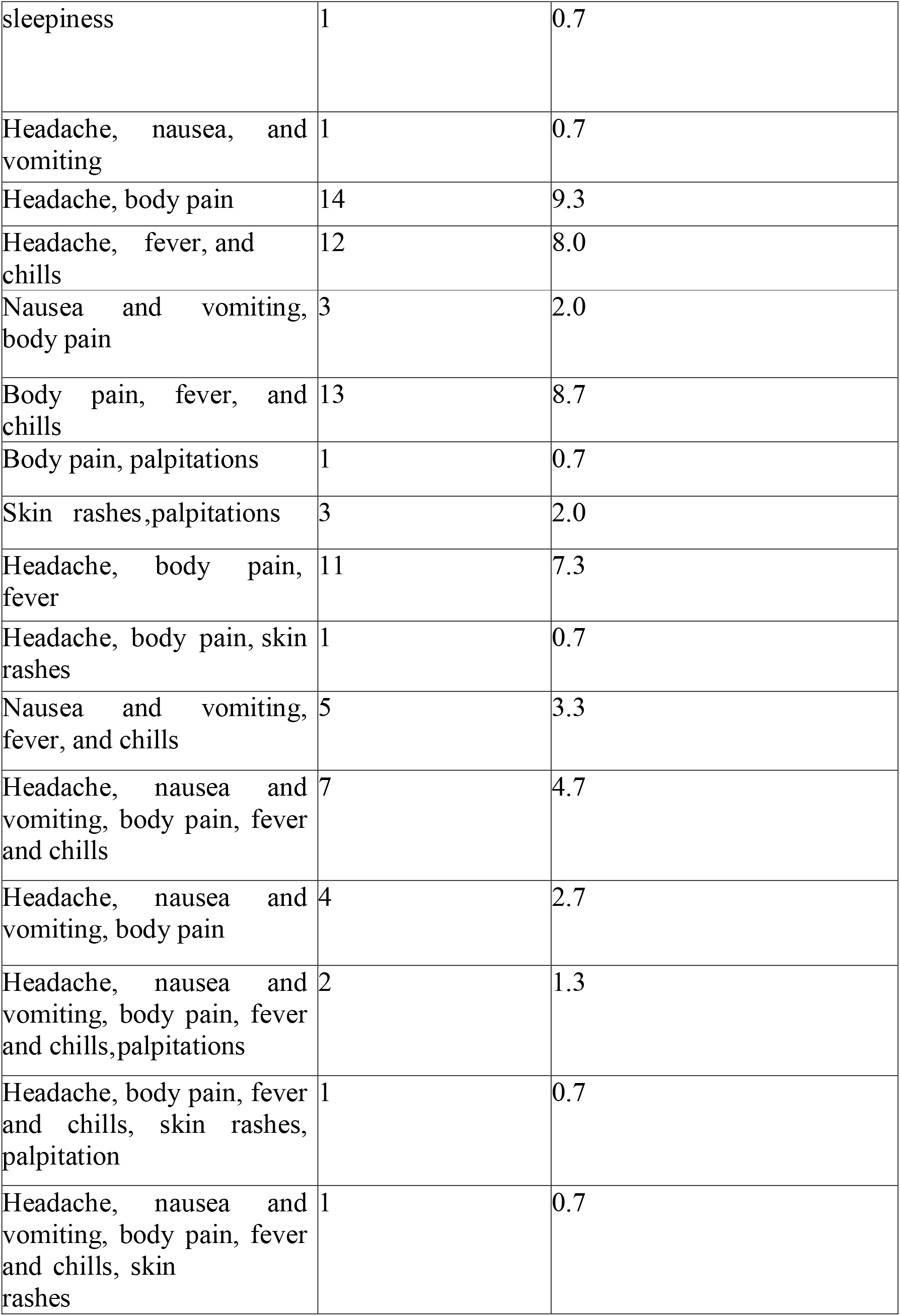
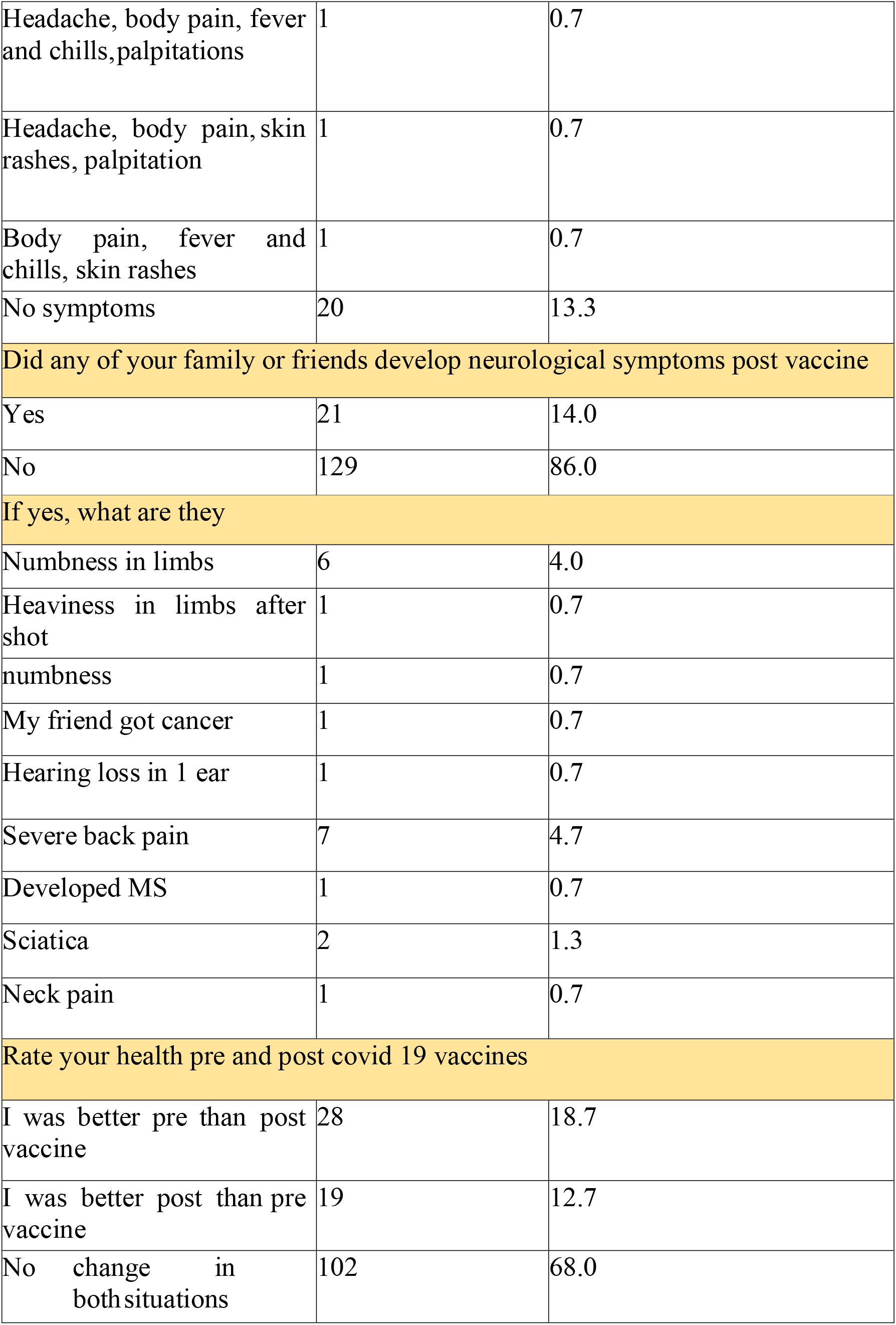
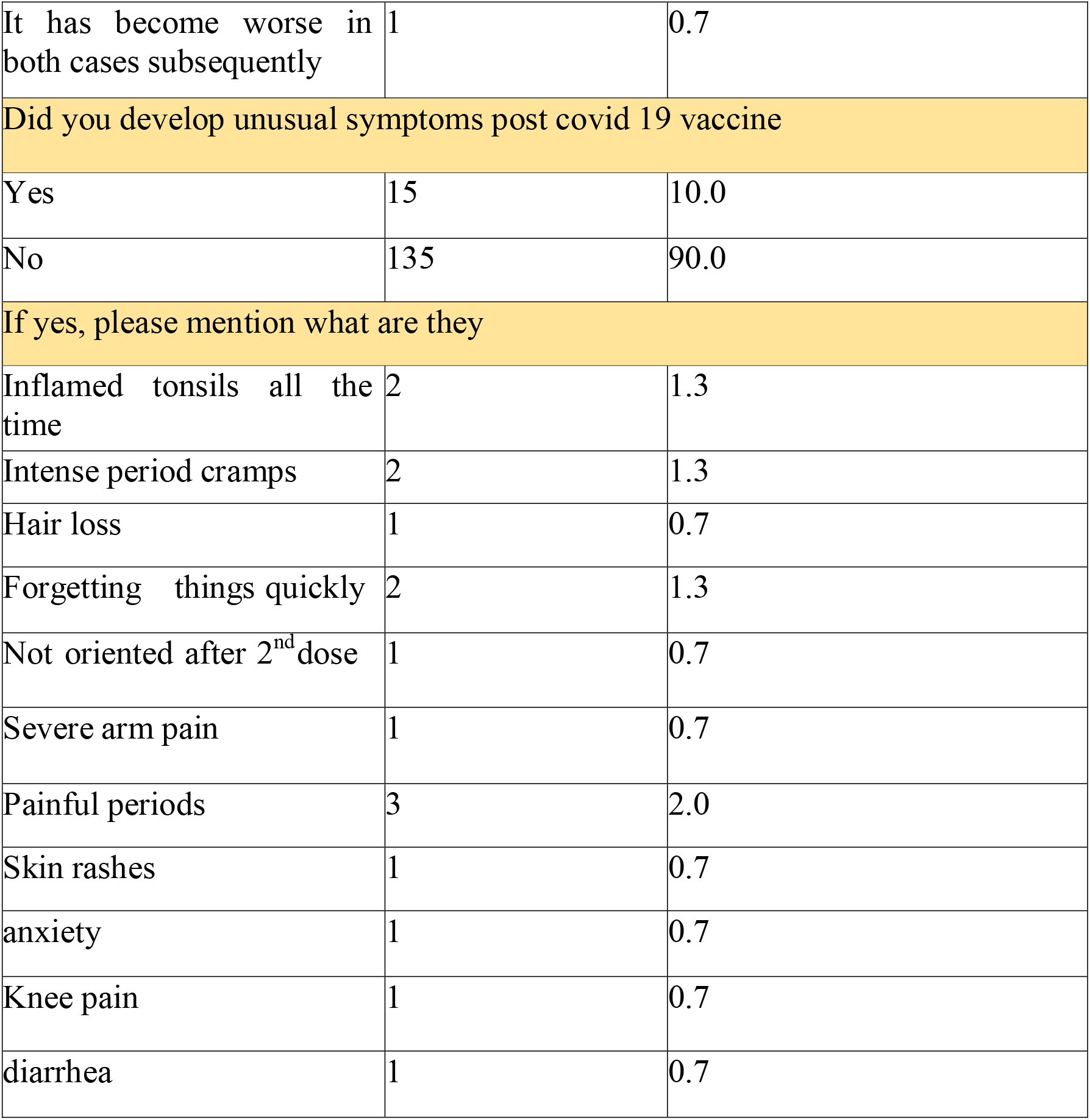
Clinical manifestation post vaccine

| **Clinical manifestation post vaccine** |  |  |
| --- | --- | --- |
| **Clinical symptoms post 1<sup>st</sup> dose of vaccine** |  |  |
|  | Frequency | Percent |
| Headache | 13 | 8.7 |
| Body pain | 29 | 19.3 |
| Fever, chills | 7 | 4.7 |
| Headache, nausea | 4 | 2.7 |
| Headache, body pain | 16 | 10.7 |
| Headache, fever, and chills | 9 | 6.0 |
| Nausea, body pain | 4 | 2.7 |
| Nausea, palpitation | 1 | 0.7 |
| Body pain, fever, and chills | 14 | 9.3 |
| Fever and chills, skin rashes | 1 | 0.7 |
| Skin rashes, palpitations | 2 | 1.3 |
| Headache, nausea and vomiting, body pain, palpitation | 1 | 0.7 |
| Headache, nausea and vomiting, body pain, fever and chills, skin rashes | 1 | 0.7 |
| Nausea and vomiting, body pain, fever, and chills | 2 | 1.3 |
| Headache, nausea and vomiting, body pain, fever, and chills | 5 | 3.3 |
| Headache, body pain, fever, and chills | 10 | 6.7 |
| Headache, body pain, diarrhea | 1 | 0.7 |
| Headache, nausea, and vomiting, body pain | 5 | 3.3 |
| Headache, body pain, fever and chills, palpitation | 2 | 1.3 |
| Headache, nausea, and vomiting, body pain, fever and chills, skin rashes | 1 | 0.7 |
| Nausea and vomiting, body pain, fever and chills, palpitation | 1 | 0.7 |
| Headache, nausea and vomiting, body pain, fever and chills, palpitations | 2 | 1.3 |
| Headache, body pain, palpitation | 1 | 0.7 |
| Headache, nausea and vomiting, body pain, fever and chills, palpitation | 1 | 0.7 |
| Injection site pain | 1 | 0.7 |
| No symptoms | 16 | 10.7 |
| Clinical symptoms post 2<sup>nd</sup> dose of vaccine |  |  |
| headache | 15 | 10.0 |
| Nausea and vomiting | 2 | 1.3 |
| Body pain | 17 | 11.3 |
| Fever and chills | 9 | 6.0 |
| Skin rashes | 2 | 1.3 |
| palpitations | 2 | 1.3 |
| sleepiness | 1 | 0.7 |
| Headache, nausea, and vomiting | 1 | 0.7 |
| Headache, body pain | 14 | 9.3 |
| Headache, fever, and chills | 12 | 8.0 |
| Nausea and vomiting, body pain | 3 | 2.0 |
| Body pain, fever, and chills | 13 | 8.7 |
| Body pain, palpitations | 1 | 0.7 |
| Skin rashes, palpitations | 3 | 2.0 |
| Headache, body pain, fever | 11 | 7.3 |
| Headache, body pain, skin rashes | 1 | 0.7 |
| Nausea and vomiting, fever, and chills | 5 | 3.3 |
| Headache, nausea and vomiting, body pain, fever and chills | 7 | 4.7 |
| Headache, nausea and vomiting, body pain | 4 | 2.7 |
| Headache, nausea and vomiting, body pain, fever and chills, palpitations | 2 | 1.3 |
| Headache, body pain, fever and chills, skin rashes, palpitation | 1 | 0.7 |
| Headache, nausea and vomiting, body pain, fever and chills, skin rashes | 1 | 0.7 |
| Headache, body pain, fever and chills, palpitations | 1 | 0.7 |
| Headache, body pain, skin rashes, palpitation | 1 | 0.7 |
| Body pain, fever and chills, skin rashes | 1 | 0.7 |
| No symptoms | 20 | 13.3 |
| Did any of your family or friends develop neurological symptoms post vaccine |  |  |
| Yes | 21 | 14.0 |
| No | 129 | 86.0 |
| If yes, what are they |  |  |
| Numbness in limbs | 6 | 4.0 |
| Heaviness in limbs after shot | 1 | 0.7 |
| numbness | 1 | 0.7 |
| My friend got cancer | 1 | 0.7 |
| Hearing loss in 1 ear | 1 | 0.7 |
| Severe back pain | 7 | 4.7 |
| Developed MS | 1 | 0.7 |
| Sciatica | 2 | 1.3 |
| Neck pain | 1 | 0.7 |
| Rate your health pre and post covid 19 vaccines |  |  |
| I was better pre than post vaccine | 28 | 18.7 |
| I was better post than pre vaccine | 19 | 12.7 |
| No change in both situations | 102 | 68.0 |
| It has become worse in both cases subsequently | 1 | 0.7 |
| Did you develop unusual symptoms post covid 19 vaccine |  |  |
| Yes | 15 | 10.0 |
| No | 135 | 90.0 |
| If yes, please mention what are they |  |  |
| Inflamed tonsils all the time | 2 | 1.3 |
| Intense period cramps | 2 | 1.3 |
| Hair loss | 1 | 0.7 |
| Forgetting things quickly | 2 | 1.3 |
| Not oriented after 2<sup>nd</sup> dose | 1 | 0.7 |
| Severe arm pain | 1 | 0.7 |
| Painful periods | 3 | 2.0 |
| Skin rashes | 1 | 0.7 |
| anxiety | 1 | 0.7 |
| Knee pain | 1 | 0.7 |
| diarrhea | 1 | 0.7 |

Although the vaccine was considered a mandatory requirement to attend classes on campus in UAE, 102 (68%) would still take the vaccine if it was optional, and 48 (32%) would not for various reasons mentioned in Table 4, highlighting the unique perception among the physiotherapist students. The most commonly reported were “they need more time to see the effectiveness 6 (4%), unnecessary 5 (3.3%), personal choice 5(3.3%). In our study, we found that a significant number of participants showing direct unwillingness to take vaccines (32%) which may be masked by the mandatory norms for class attendance. Nevertheless, the findings were in line with previous studies conducted among various populations. A study conducted on medical students in the USA reported that 53% would be willing to participate in the covid -19 vaccine trial and 23% were unwilling [13]. As the data from this study is directly comparable to our study, we can suggest that medical students in UAE showed a higher rate of unwillingness or hesitancy to be vaccinated against Covid-19. The higher rate of willingness to take vaccines in the USA was attributed to trust in public health experts, fewer concerns about side effects, and agreement with vaccine mandates. In comparison to our study for a higher willingness rate of 68%, a study conducted on general university students in UAE reported an acceptance rate of 56.3 % where the worries regarding unforeseen problems were significantly higher (65.5%) [12]. The difference in the findings could be attributed to professional medical stream-oriented students in our study which led to higher values for vaccine acceptance and positive perception.

The important findings regarding reported clinical manifestation are presented in Table 5. The students were given multiple options to report their clinical manifestations post-vaccination for the first and seconds dose. The most frequently reported symptoms after the first dose were “body pain” reported by 29 students (19.3%) followed by headache with body pain (n=16,10.7%), whereas 16 students (10.7%) reported no symptoms. A similar pattern was noticed in the symptoms reported post seconds dose where 17 students (11.3%) reported body pain, 15 (10%) headache, and 17 (11.3%) headache with body pain, while 20 (13.3%) reported no symptoms. In September 2021, a paper published concluded that delayed onset of headaches post covid 19 vaccine can be a red flag related to the development of cerebral venous thrombosis and individuals reporting suchsymptoms should undergo further clinical assessment [21]. The results also indicated that 21 (14%) students had friends and family developed neuromuscular symptoms post-vaccine while 129 (86%) reported no symptoms. The most commonly reported symptoms were severe back pain (7,4.7%), numbness in the limb (6, 4%), and sciatica (2,1.3%). Though these findings have higher clinical significance, there is no comparable data to support the evidence currently. We recommend that these findings should be taken seriously as they may impact the professional and personal life of growing professionals. In context to rating the health pre and post-Covid-19 vaccination, 102 students (68%) rated their health as having no change after both doses, 28 (18.7%) stated that “I was better pre than post-vaccine”, 19 (12.7%) rated as “I was better to post than pre-vaccine” and 1(0.7%) it has become worse in both cases subsequently. It was positive to report that the majority of participants found no change in health status, however, significant participants were affected post vaccination which reflects the need to explore the reasons associated with vaccination. In continuation to such findings, students were asked if they did develop any unusual symptoms post-vaccine and if yes to mention what they were. It is important to mention that 15 students (10%) answered “Yes” and 135 (90%) answered “No”. Among the respondents for “Yes”, the most common unusual symptoms were painful periods 3 (2%), inflamed tonsils all the time 2(1.3%), intense period cramps 2 (1.3%), and less common were hair loss, severe arm pain, skin rashes, anxiety and forgetting things quickly. These findings were very new to highlight among university students and have not been reported in the previous literature. It should be noted that the clinical manifestation reported here was of prime concern which could have again affected the student’s regular and professional work significantly. Also, it could impose extra mental stress which could have affected their contribution to the medical care of patients when posted for clinical exposure.

Although the findings of our students were not comparable to previous studies due to the lack of similar findings and the novelty of the research area, there are a few important studies to report. The study conducted by Shahwan et al. (2022) suggested that there was a higher degree of variability toward knowledge of Covid-19 among students and misconceptions needed to be addressed [12]. To enhance Covid-19 vaccination uptake in the country and worldwide, health education targeting diverse sociodemographic categories should be prioritized. Similarly, the study conducted by Qiao et al. (2022) among a larger population of students, found that the perceived severity of Covid-19 was positively associated with vaccine acceptance [22]. Higher levels of risk exposure and negative attitude toward general vaccination were associated with low vaccine acceptance. They suggested the need for tailored education messages for college students to emphasize the severity of Covid-19. Also, it was important to address the concerns of side effects of general vaccines by dispelling the misconception and targeting the most vulnerable subgroups who reported high levels of risk exposures while showing low intention to take the vaccine. Whereas the study conducted by Lucia et al. (2021) among medical students found that nearly all participants had positive attitudes toward vaccines and agreed that they would likely be exposed to Covid-19 [13]. Among the general population, the study conducted by Maria et al. (2021) suggested that vaccine hesitancy was present in the study population with 32.6% being unsure and 15.6% declaring that they were not willing to take the vaccine [23]. Females were more likely to be unsure. Lack of vaccine safety was the main reason for the unwillingness to take the vaccine. Predictors for willingness to take the vaccine were: i) The beliefs that the Covid-19 vaccine will protect the health of the people who take it; ii) the advice of health professionals regarding the effectiveness of the Covid-19 vaccine; iii) Having taken the influenza vaccine last year and iv) Encouraging their elderly parents to take the vaccine.

It was evident from the findings of this study that healthcare care professionals such as physiotherapy students had unique experiences towards Covid-19 pandemic with specific beliefs, perceptions, and clinical manifestations which could have affected their regular and professional work, lifestyle changes, and social life. It was also important to highlight various reasons associated with their mental and physical health during the pandemic. The findings of the study could be used to counsel the physiotherapy students or similar cohort to address their concerns and fears they might be experiencing as well as to direct them towards the right sources of information before any new roles and responsibilities which could be of first-lifetime experience such as Covid-19 pandemic. Certain findings of the study require proper concern and medical attention especially a sudden onset of numbness and severe low back pain that may affect their quality of life in all domains including psychosocial. Also, these findings suggest the impact of clinical manifestations associated with vaccinations which could have affected their academics and professional skills and thus proper preventive measures should be taken to counter such concerns with necessary facilities and infrastructure.

### Strength

This study is considered the first to explore the beliefs and perceptions of medical students specifically physiotherapy students in the UAE regarding covid 19 vaccine, as well as the clinical manifestations post covid 19 vaccine has been explored in this study.

### Limitations

The participants consisted of predominantly females which may have affected the findings of the study due to the gender variations in responses. The population was heterogeneous in terms of race, culture, and socioeconomic status, which could have affected the findings of the study and could not be controlled. The survey was collected through google forms, resulting in challenges to get responses which lead to 70 percent of desired sample size.

## Conclusion

Findings of the study concluded that a significant number of physiotherapy students believed that covid 19 was not safe and showed resistance toward acceptance. They perceived that post covid 19 vaccination was associated with reduced physical activity due to its effect on physical and mental health. Necessary steps need to be taken to counsel the students’ groups for better preparation toward their readiness and participation in the healthcare system for the future.

## Data Availability

All data produced in the present study are available upon reasonable request to the authors. The data underlying the results presented in the study are available from the corresponding author. Please contact us via mail after publication.

